# Neutralizing efficacy of vaccines against the SARS-CoV-2 Mu variant

**DOI:** 10.1101/2021.09.23.21264014

**Authors:** Kei Miyakawa, Sundararaj Stanleyraj Jeremiah, Hideaki Kato, Akihide Ryo

## Abstract

The rise of mutant strains of SARS-CoV-2 poses an additional problem to the existing pandemic of COVID-19. There are rising concerns about the Mu variant which can escape humoral immunity acquired from infections from previous strains or vaccines. We examined the neutralizing efficacy of the BNT162b2 mRNA vaccine against the Mu variant and report that the vaccine has 76% neutralizing effectiveness against the Mu compared to 96% with the original strain. We also show that Mu, similar to the Delta variant, causes cell-to-cell fusion which can be an additional factor for the variant to escape vaccine-mediated humoral immunity. Despite the rise in vaccine escape strains, the vaccine still possesses adequate ability to neutralize majority of the mutants.

## Main text

The rapid and nearly unrestricted global spread of COVID-19 has ensued the evolution of various mutants of SARS-CoV-2. With vaccines being the principal effective modality to curtail the pandemic, we are left with no option than to use them and brace for the rise of immune escape mutants that can evolve due to the selection pressure exerted. Based on clinical and epidemiological significance, the WHO has identified four variants of concern (VOC); Alpha, Beta, Gamma and Delta, and five variants of interest (VOI); Eta, Iota, Kappa, Lambda and Mu [1]. Despite Delta being the principal mutant responsible for majority of the infections at present, the new kid on the block; Mu has caused a significant amount of commotion due to its higher propensity for immune escape [2]. In this background, we wanted to evaluate the significance of this alarm raised by Mu with respect to the efficacy of vaccine derived neutralizing antibodies (nAb) and that of the dual antibody cocktail therapy against this variant.

We performed the VLP (virus-like particle)-based rapid neutralization test (hiVNT) [3, 4] on post-vaccination sera collected from individuals one week after administration of the second dose of the BNT162b2 mRNA vaccine. The serum dilution factor that inhibits VLP entry by half (NT_50_) was assessed to demonstrate the neutralizing activity of these sera against the variants Mu, Alpha, Beta, Gamma, Delta and Lambda. The median neutralizing efficacy for all the variants was above the effective threshold (NT_50_=20) in all the tested sera suggesting that the vaccine derived nAbs can neutralize majority of all the variants including Mu (Figure 1A, S1A). Previously infected individuals showed prominently higher NT_50_ values for all variants after vaccination (Figure S2).

**Figure 1.**
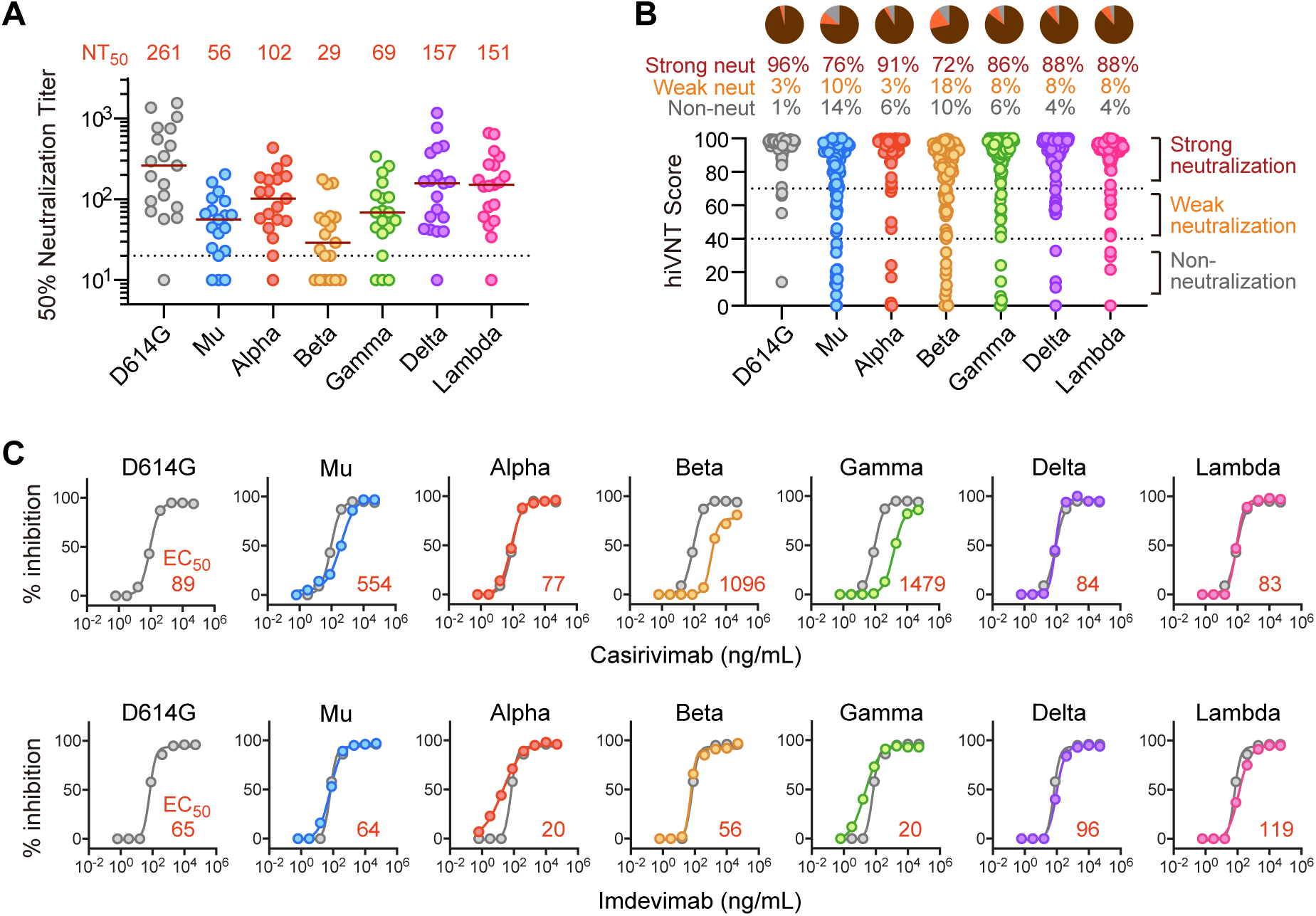
Neutralization of SARS-CoV-2 Mu variant by vaccine sera and monoclonal antibodies. **(A)** Neutralizing activity of BNT162b2 post-vaccination sera (1 week after second dose) against each variant (n=19). Serum dilutions showing 50% inhibition of infection (NT_50_) were determined by quantitative hiVNT. The dotted line indicates the cut-off threshold of this assay (NT_50_=20). The red lines indicate the median NT_50_ and the values of which are displayed as red numerals above each parameter in the graph. Note that this assay correlates well with the assay using the authentic virus (see Figure S1A). **(B)** Positive rates for neutralizing antibody determined by qualitative hiVNT. BNT162b2 post-vaccination sera (1 week after second dose) were used (n=105). The percentage of inhibition of viral infection by 20-fold dilution of serum is shown in the scatter plot as hiVNT score. hiVNT score below 40 (equivalent to pvNT_50_ < 50) is defined as nAb-negative serum, 40-70 (equivalent to pvNT_50_ > 50 but < 200) as weakly neutralizing serum, and above 70 (equivalent to pvNT_50_ > 200) as strongly neutralizing serum. The percentage of each serum is shown in the pie chart. Note that the qualitative hiVNT provides semi-quantitative results (see Figure S2B for the description of the accuracy of this definition). **(C)** Neutralization curve of each mutant strain by two monoclonal antibodies (Casirivimab and Imdevimab). The red numbers indicate the 50% effective concentration (EC_50_, ng/mL). For comparison, the neutralization curve for each mutant is superimposed on that of the D614G control (gray line). Since these nAb medicines are treated as a cocktail, they are considered effective if the EC_50_ of either antibody is equivalent to or lower than that of the D614G control.

We then wanted to grade the efficacy of the nAb response in post vaccinated sera against the different variants. For this we used pvNT_50_ (Serum dilution factor that inhibits HIV-based pseudovirus infection by half) of 40 as the lower threshold and pvNT_50_ of 200 as the higher threshold. This is based on a recent report that the pvNT_50_ in sera of individuals with vaccine-breakthrough infections was around 200 [5]. Samples that fell below the lower threshold were deemed negative for neutralizing activity, while those in between the lower and higher threshold were considered to possess weak neutralizing activity and those above the higher threshold reflected strong neutralizing activity (Figure 1B, S1B). Strong neutralization of all variants was observed in majority of the sera ranging from the highest of 96% in D614G to the lowest 72% in Beta variant. Mu showed a pattern similar to Beta with 76% of the samples strongly neutralized. The proportion of sera that did not neutralize was much lower when compared to those possessing neutralizing efficacy for each variant. The highest occurrence of nAb escape (including weak and non-neutralizing activity) was noted with Beta (28%) followed by Mu (24%).

We then evaluated the efficacy of the dual antibody cocktail against these variants and found that all the tested variants including Mu were neutralized by at least one of the two antibodies in the cocktail (Figure 1C). We further demonstrate that the Mu variant can also cause cell-cell fusion (Figure S3) like Delta variant, which is highly likely to promote the viral resistance to nAbs [6].

With rise of the Mu variant, there has been a panic concerning the efficacy of the currently available vaccines. Our results show that the vaccine derived nAbs and the antibody cocktail still possess adequate neutralization efficacy against this variant. We observed this effect in sera of vaccine recipients shortly after the receipt of the second dose when the nAbs are supposed to be at peak levels. As vaccine derived nAbs can wane over time, follow-up studies are needed to assess the persistence of nAbs against the Mu variant. By possessing the property of cell fusion like the Delta variant, but with a higher proportion of escape from the vaccine than the latter, Mu could especially be a problem if it would replace the Delta as the most predominant variant. Despite this, the current vaccines and antibody cocktail would still work for the majority, albeit at a slightly lowered efficacy than currently observed with the Delta variant.

## Data Availability

The datasets presented in this article are not readily available because it is difficult to ensure the de-identification of data. However, they can be available from the corresponding authors on reasonable request.

## Acknowledgement

We acknowledge all medical staff involved in the study. We thank Kenji Yoshihara and Kazuo Horikawa for their technical assistance. This study was supported by AMED grants (JP20he0522001, JP21fk0108104) to AR.

## Conflict of interests

Authors have no conflicts of interest directly relevant to the content of this article.

## Author contributions

KM designed and performed the research, analyzed the data, and wrote the manuscript; SSJ analyzed the data and wrote the manuscript; HK collected the specimens; AR directed the research, analyzed the data, and wrote the manuscript.

## Supplementary Appendix

**Supplementary Figure S1.**
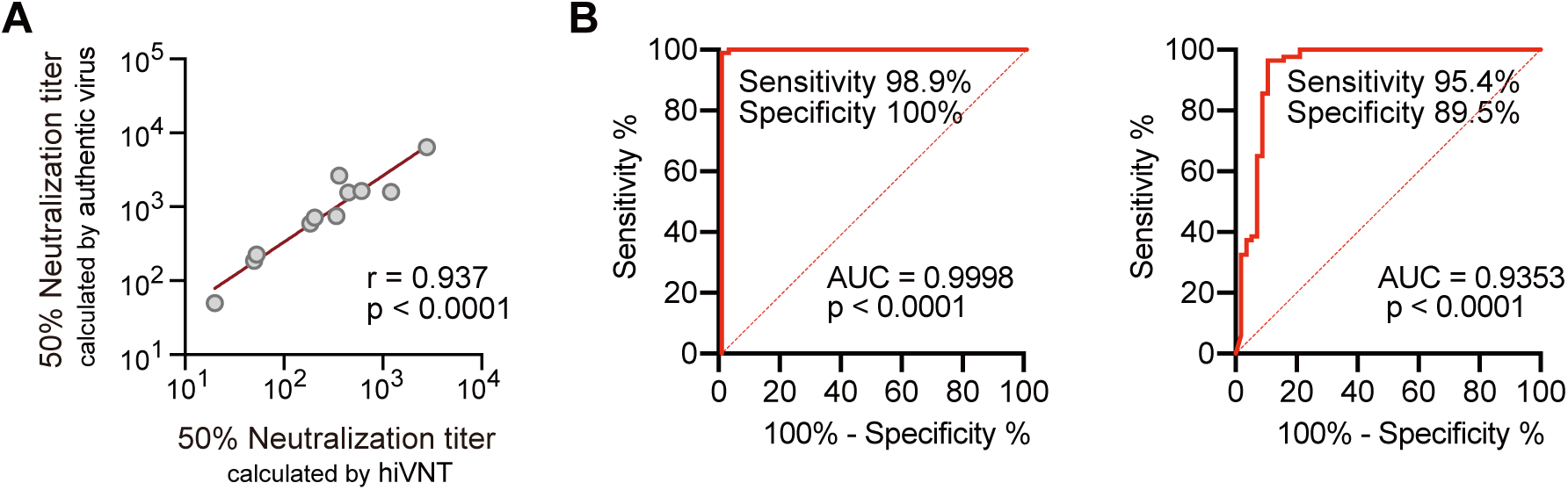
Accuracy of hiVNT used in this study. **(A)** Correlation between the quantitative hiVNT and the standard neutralization assay using authentic virus. Eleven sera were plotted as serum dilutions showing 50% inhibition of infection by each method. **(B)** Accuracy of qualitative hiVNT scoring and neutralization activity. On the left panel shows the receiver operating characteristic (ROC) curve when a hiVNT score of 40 is defined as a nAb negative (pvNT_50_ < 50) in the pseudovirus neutralization test. On the right panel shows the ROC curve when a hiVNT score of 70 or higher is defined as a serum with strong neutralizing activity (NT_50_ > 200). Although the qualitative hiVNT method measures only 20-fold dilution of serum, it still provides semi-quantitative results [note that Area under curve (AUC) >0.90 in both cases].

**Supplementary Figure S2.**
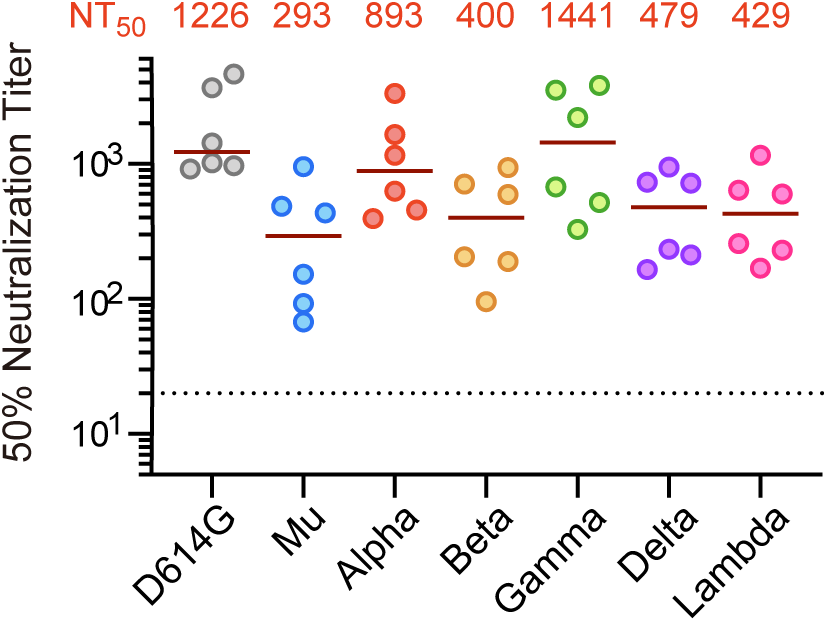
Neutralization of SARS-CoV-2 Mu variant by vaccine sera in previously infected individuals. Neutralizing activity of sera after BNT162b2 vaccination (1 week after the second dose) against each variant in previously infected individuals (n=6). The red line and numbers in the graph indicate the median values. Serum dilutions that inhibit infection by 50% were determined by quantitative hiVNT. The dotted line indicates the threshold value in this assay.

**Supplementary Figure S3.**
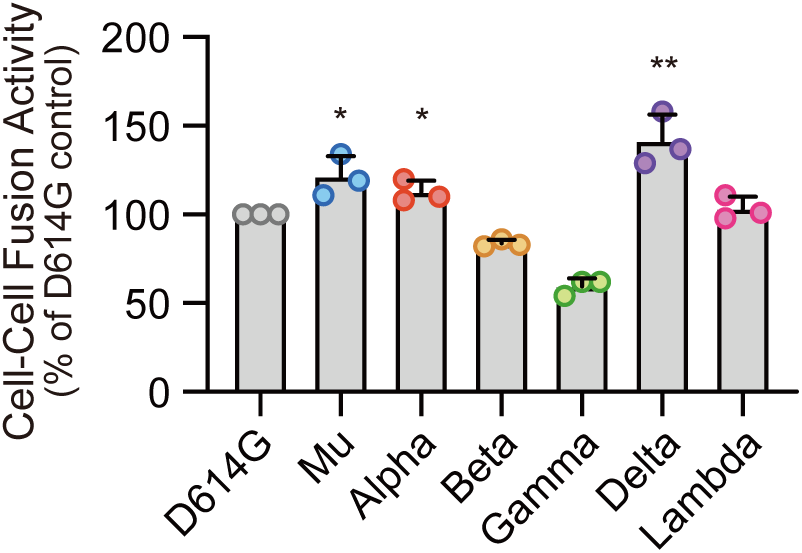
Cell fusion activity of SARS-CoV-2 variant. Cell-to-cell fusion activity of indicated variants. Assay was carried out using a split luciferase system. HEK293-donor cells (expressing SARS-CoV-2 spike and HiBiT) and HEK293-acceptor cells (expressing LgBiT and ACE2) were cocultured in 1:1 ratio. After 3 hours, the luciferase activity derived from cell fusion was measured. \**P* < 0.05, ** *P* < 0.01. Higher luciferase signal denotes higher occurrence of cell-to-cell fusion.

## Supplementary Materials

### Ethics statement

This study was approved by Yokohama City University Certified Institutional Review Board (Reference No. B210300001), and the protocols used in the study were approved by the ethics committee. Written informed consent was obtained from all the participants.

### Rapid neutralization test (hiVNT)

hiVNT was performed as previously described [3, 4]. Briefly, target cells seeded in 96-well plates were inoculated with 50 µL of HiBiT-tagged virus-like particles (hiVLPs) containing diluted serum (1:20–1:43,740 dilution for quantitative assay; 1:20 dilution for qualitative assay). At 3 hours after inoculation, intracellular luciferase activity is measured. The dilution factor of serum that resulted in a 50% reduction in luminescence compared with the non-serum control was set as the NT_50_. We calculated NT_50_ using the curve-fitting tool (ImageJ, NIH). When serum had no observable neutralizing activity to interpolate NT_50_, it was assigned an NT_50_ of 10. All samples were assayed in duplicate.

In qualitative assay, the hiVNT score (percentage of luminescence signal inhibition) was calculated as follows:

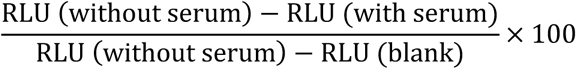

Alternatively, cells were inoculated with 50 µL of hiVLPs containing diluted antibody (final concentration of 0.64–50,000 ng/mL). The concentration of antibody that resulted in a 50% reduction in luminescence compared with the non-antibody control was set as EC_50_.

## Notes

### Competing Interest Statement

The authors have declared no competing interest.

